# Kinetics of viral clearance and antibody production across age groups in SARS-CoV-2 infected children

**DOI:** 10.1101/2020.08.06.20162446

**Authors:** Burak Bahar, Cyril Jacquot, Yunchuan D. Mo, Roberta L. DeBiasi, Joseph Campos, Meghan Delaney

## Abstract

**Objectives:** To improve understanding of transition from viral infection to viral clearance, and antibody response in pediatric patients with SARS-CoV-2 infection.

**Study design:** This retrospective analysis of children tested for SARS-CoV-2 by RT-PCR and IgG antibody at a quaternary-care, free-standing pediatric hospital between March 13^th^, 2020 to June 21^st^, 2020 included 6369 patients who underwent PCR testing and 215 patients who underwent antibody testing. During the initial study period, testing focused primarily on symptomatic children; the later study period included asymptomatic patients who underwent testing as preadmission or preprocedural screening. We report the proportion of positive and negative tests, time to viral clearance, and time to seropositivity.

**Results:** The rate of positivity varied over time due to viral circulation in the community and transition from targeted testing of symptomatic patients to more universal screening of hospitalized patients. Median duration of viral shedding (RT-PCR positivity) was 19.5 days and RT-PCR negativity from positivity was 25 days. Of note, patients aged 6 to 15 years demonstrated a longer period of RT-PCR negativity from positivity, compared to patients aged 16 to 22 years (median=32 versus 18 days, p=0.015). Median time to seropositivity from RT-PCR positivity was 18 days while median time to reach adequate levels of neutralizing antibodies (defined as equivalent to 160 titer) was 36 days.

**Conclusions:** The majority of patients demonstrated a prolonged period of viral shedding after infection with SARS CoV-2. Whether this correlates with persistent infectivity is unknown. Only 17 of 33 patients demonstrated neutralizing antibodies, suggesting that some patients may not mount significant immune responses to infection. It remains unknown if IgG antibody production correlates with immunity and how long measurable antibodies persist and protect against future infection.

## Introduction

In December 2019, a series of severe acute respiratory syndrome (SARS) cases caused by a novel strain of coronavirus (CoV), believed to have originated from bats, was described in Wuhan, Hubei province of China.^1^ The pathogen demonstrated a significant degree of genetic homology to the SARS virus from the 2002-2004 outbreak. The virus strain was dubbed SARS-CoV-2 and the disease was named coronavirus disease 2019 (COVID-19). A pandemic was declared by the World Health Organization (WHO) on March 11, 2020 after the number of affected cases had increased significantly and the disease was observed in more than 100 countries.^2^

SARS-CoV-2 has similar structural proteins present in other CoVs, consisting of S (spike), E (envelope), M (membrane), and N (nucleocapsid) components.^3, 4^ Of these glycoproteins, virus-cell fusion of SARS-CoV-2 is mediated by the trimeric structure of the two functional subunits of the S protein, namely S1 and S2, after binding to angiotensin-converting enzyme 2 (ACE2).^5^ Antibodies formed against the receptor binding domain (RBD) on the S1 subunit have the potential to neutralize SARS-CoV-2 by disabling virus-ACE2 binding and endocytosis.^6, 7^ In addition, the competitive RBD binding capacity of these antibodies versus ACE2 correlates with neutralizing activity. ^6^

Dong *et al*. reported the first epidemiological results on 728 children with laboratory-confirmed COVID-19 from China.^8^ The authors found that the median age was seven years, greater than 90% of the cases had a disease spectrum ranging from asymptomatic to moderate disease, and the proportion of severe and critical cases increased with age.^8^ Similar findings have subsequently been reported from Europe, the middle east, and the US.^9–12^

Although there is emerging data regarding timing of viral clearance and immunological response in adults with COVID-19^13^, there is a scarcity of data in the pediatric population. Furthermore, a lack of knowledge regarding factors affecting time-to-seropositivity is observed in both pediatric and adult patients.^14^ We report viral and antibody testing results from our pediatric patient population in order to contribute to a better understanding of timing of viral clearance and antibody production in children with COVID-19.

## Methods

From March 13^th^, 2020 to June 21^st^, 2020, all patients presenting to our institution (Children’s National Hospital, Washington, DC) who were aged ≤ 22 years and had been tested for SARS-CoV-2 by reverse transcription polymerase chain reaction (RT-PCR) were included in the study. Data were extracted from our laboratory information systems data warehouse (Sunquest Information Systems Database [Tucson, AZ, USA] and Vertica Analytics Platform [Cambridge, MA]) using Viewics Analytics Platform (Roche, Indianapolis, IN) with Structured Query Language (SQL) queries. Extracts were merged into CSV files and imported into RStudio IDE (Boston, MA). In addition to the RT-PCR results, qualitative and quantitative serologic testing results, age and sex were also included in the data extracts. Age stratifications were defined as 0 to 5 years, 6 to 15 years and 16 to 22 years (see supplemental figure S1 for kernel density estimate of age).

Nasopharyngeal samples were collected from patients. Samples were transferred to the laboratory in viral transport medium as recommended by the manufacturer or in liquid Amies medium as validated by the laboratory. For detection of the virus by RT-PCR, four systems were utilized due to high testing volume, which were (1) GenMark ePlex SARS-CoV-2 Test, (2) DiaSorin Molecular Simplexa™ COVID-19 Direct Assay System, (3) Cepheid Xpert^®^ Xpress-SARS-CoV-2 and (4) Seegene Allplex™ 2019-nCoV RT-PCR Assay. All were validated in our laboratory and considered equivalent for patient testing. Viral clearance was deemed as RT-PCR negativity.

The GenMark ePlex SARS-CoV-2 Test is an automated qualitative nucleic acid test that aids in the detection of SARS-CoV-2 and diagnosis of COVID-19 using The True Sample-to-Answer Solution™ ePlex instrument. The test is based on nucleic acid amplification technology and each test cartridge includes all reagents needed to extract, amplify and detect SARS-CoV-2 RNA in nasopharyngeal swab samples.^15^

The DiaSorin Molecular Simplexa™ COVID-19 Direct Assay System is a real-time RT-PCR system that enables the direct amplification of Coronavirus SARS-CoV-2 RNA from nasopharyngeal swabs. Fluorescent probes are used together with corresponding forward and reverse primers to amplify SARS-CoV-2 viral RNA and internal control RNA. The assay targets two different regions of the SARS-CoV-2 genome, ORF1ab and S gene. The S gene encodes the spike glycoprotein of the SARS-CoV-2 and is also targeted to specifically detect the presence of SARS-CoV-2. The ORF1ab region encodes well-conserved non-structural proteins and therefore is less susceptible to recombination. An RNA internal control is used to detect RT-PCR failure and/or inhibition.^15^

The Cepheid Xpert^®^ Xpress-SARS-CoV-2 is a real-time RT-PCR assay designed to detect SARS-CoV-2 nucleic acids from upper respiratory samples. Specific molecular targets include the E and N2 genes. The GeneXpert platform utilizes a closed cartridge system which does not require separate extraction or processing steps prior to introduction of the sample and can yield rapid results within 30 minutes. Each cartridge contains internal sample processing and probe check controls.^15^

The Seegene Allplex™ 2019-nCoV RT-PCR Assay detects SARS-CoV-2 nucleic acids in upper respiratory samples. The probes are designed to target the E, N, and RdRp (RNA-dependent RNA polymerase) genes. A separate nucleic acid extraction step is required prior to amplification via real-time PCR. High throughput analysis is enabled through use of 96-well plates. Internal positive and negative controls are used to confirm the validity of each PCR run.^15^

Antibody detection was performed from serum or plasma samples, collected in appropriate separator tubes, using DiaSorin Liaison XL SARS-CoV-2 IgG S1/S2 assay (DiaSorin, Saluggia, Italy). We validated the analytical and clinical performance of this assay with similar outcomes as other researchers who previously reported satisfactory results.^16, 17^ The test is based on chemiluminescent detection of antibodies against S1 and S2 glycoproteins of the virus using magnetic beads. Seropositivity was defined at presence of anti-SARS-CoV-2 IgG antibodies ≥ 15 absorbance units per milliliters (AU/mL), as recommended by the manufacturer. In addition to the qualitative results, quantitative test results were also included to reflect the amount of circulating IgG antibodies in patient samples at the time of blood draw. As demonstrated by the manufacturer, antibody results ≥ 80 AU/mL were comparable to a titer of 160 by plaque reduction neutralization testing (Liaison SARS-CoV-2 S1/S2 IgG (REF 311450)). According to these criteria, results were grouped as ‘adequate for neutralization’ and ‘not adequate for neutralization’, based on the 80 AU/mL threshold. In addition to routine testing ordered by clinical providers (n=194), we retrieved available leftover serum or plasma samples from patients who underwent RT-PCR testing but were not tested for antibodies (n=19). Serologic testing was also performed on these samples and their results were included in the present study.

Statistical analyses were performed using R software (R foundation for Statistical Computing, version 4.0.0). Survival (version 3.2-3) and SurvMiner (version 0.4.7) packages were utilized for nonparametric estimation of time to event function. Interval censoring was present since timing of symptom onset was not included as a variable (left censoring) and patients were tested at the discretion of the healthcare provider and exact event times could not be obtained (right censoring). Follow-up time for patients included RT-PCR positivity duration (censored time), initial RT-PCR positivity to RT-PCR negativity (event time), initial RT-PCR positivity to seronegativity (censored time) and initial RT-PCR positivity to seropositivity (event time). These time points were utilized in event probability estimates using the Kaplan-Meier method and Peto & Peto modification of the Gehan-Wilcoxon test for comparison of groups.

Statistical significance was defined at a p-value of <0.05. Bonferroni adjustments were made to p-values for pairwise comparisons. Results were expressed in mean ± standard deviation (SD), median and 1^st^ and 3^rd^ interquartile ranges (IQR) or minimum (min) and maximum (max), and with 95 percent confidence intervals (95% CI), as appropriate.

This project was undertaken as a quality improvement initiative at Children’s National Hospital and therefore does not constitute human research. As such, it was not under the oversight of the institutional review board. This manuscript was evaluated and approved by the institutional publication review committee.

## Results

The total number of RT-PCR tests performed over the 100-day period was 7958 with 641 positive and 7317 negative test results (Figure 1). Figure 2 shows the number of patients at each stage of the study. Five hundred and ninety-two (592) patients tested positive with a median test of one per patient (max = 6). Five thousand seven hundred and seventy-seven (5777) patients tested negative with a median test of 1 per patient (min = 1, max = 15). Two hundred and thirty-eight (238) serology tests were performed with 69 positive and 169 negative test results. Overall, 58 patients tested positive with a median of one test per patient (max = 2) and 157 patients tested negative with a median test of one per patient (max = 5).

**Figure 1:**
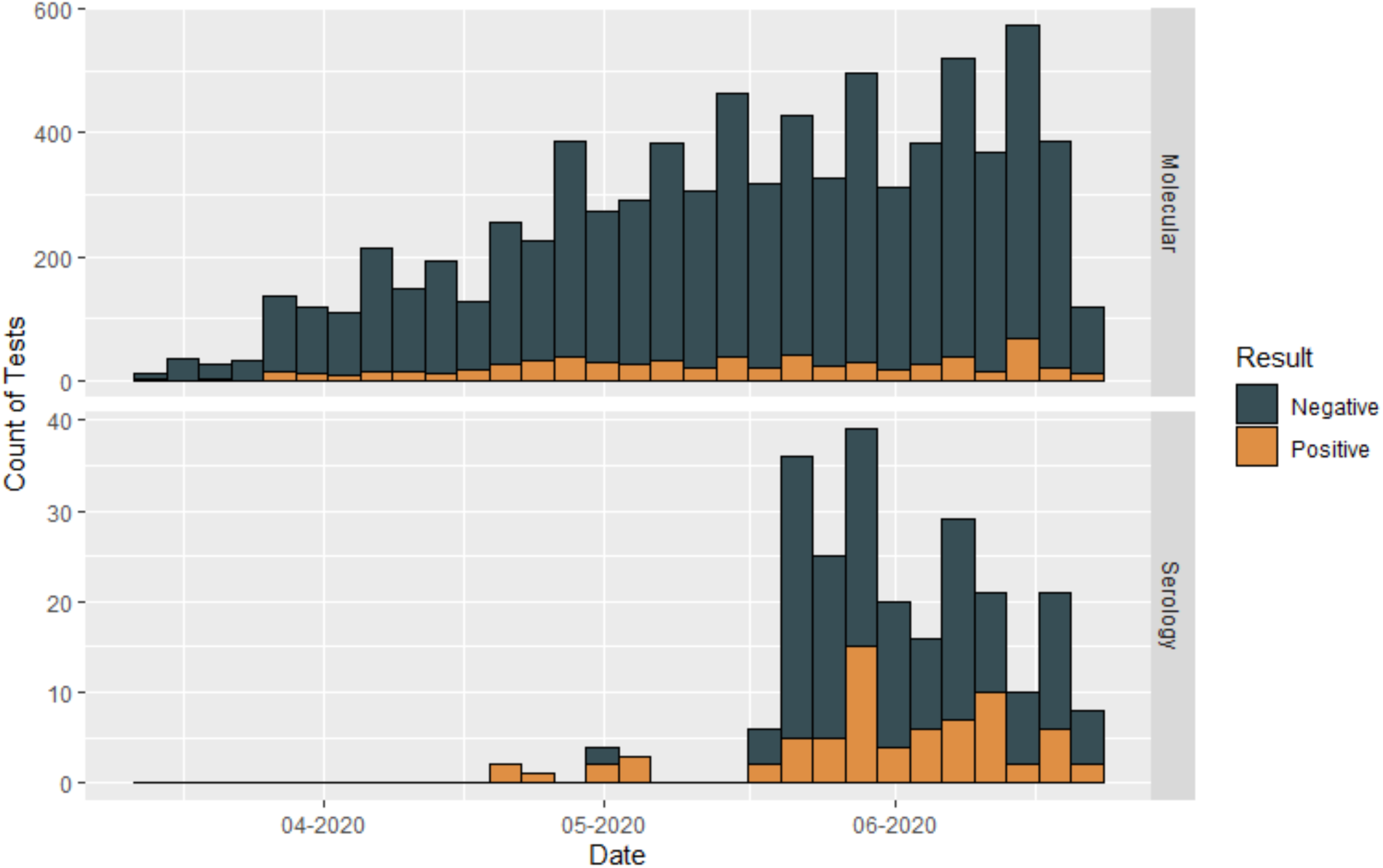
Number of molecular (RT-PCR) and serology tests performed during the study period

**Figure 2:**
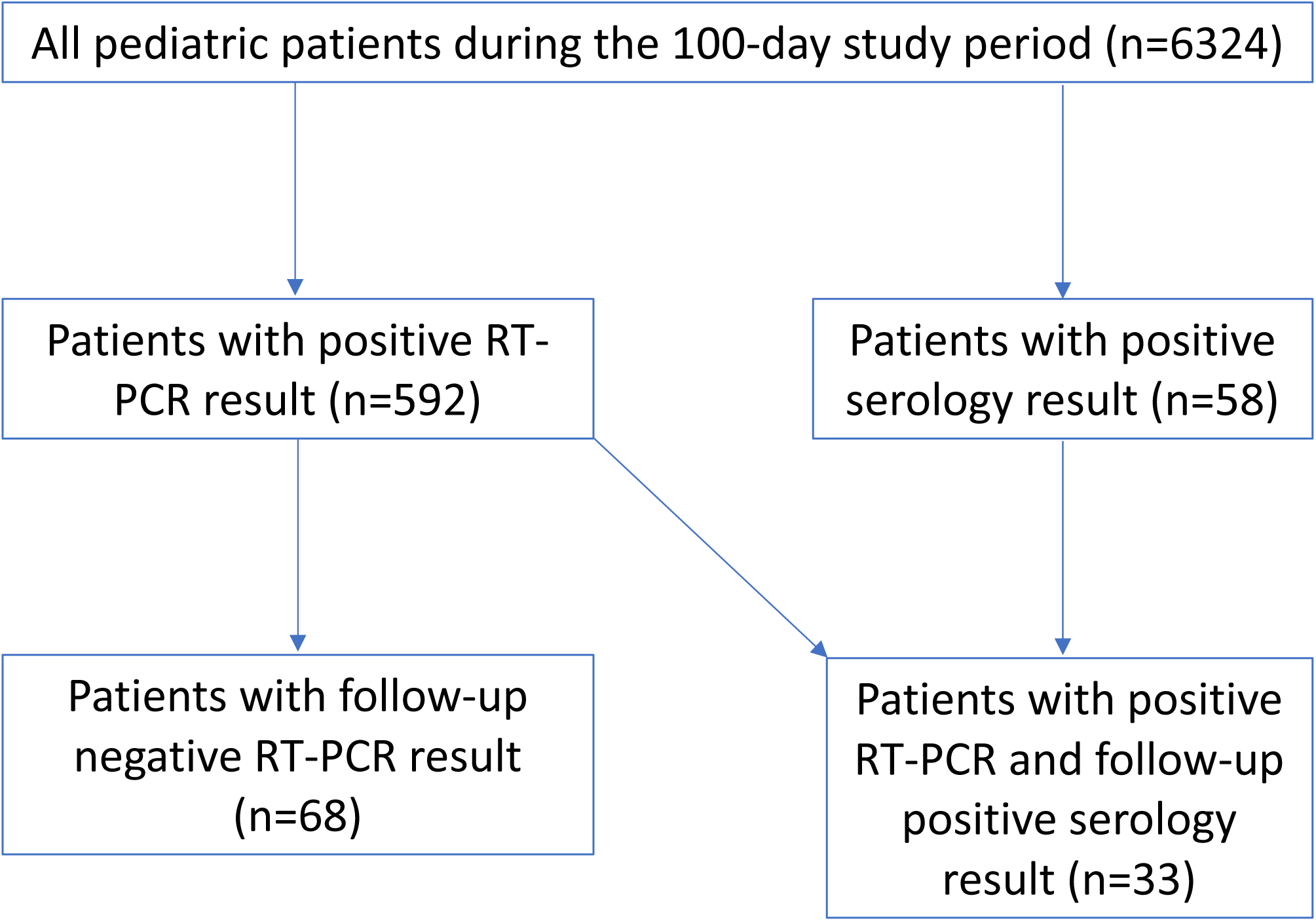
Participant flow in the study

Sixty-eight (68) patients had more than one molecular test performed. The median duration of viral shedding (RT-PCR positivity) was 19.5 days (IQR = 12-39) with 10 patients demonstrating a duration greater than 30 days (max = 62 days). The median time to RT-PCR negativity from RT-PCR positivity was 25 days (95% CI=22-34) (Figure 3a). No difference was found between females (median = 26 days) and males (median = 25 days) for time to RT-PCR negativity (χ^2^=0, p=1) (Figure 3b), however, statistical significance was found between age groups (χ^2^=7.4, p=0.02). Patients aged 6 to 15 years took longer to achieve RT-PCR negativity (median = 32 days) compared to those who were 16 to 22 years of age (median=18 days) (p=0.015) (Figure 3c). Patients in the 0 to 5 year age group required a median of 22 days to become RT-PCR negative but pairwise comparisons of this group with other groups were not significant (vs 6 to 15 years: p=0.76; vs 16 to 22 years: p=0.52). After adjustment for sex, time to RT-PCR was found to be longer only for females (n = 10, median = 44 days) in this age group since males (n = 19) in the same age cohort demonstrated a median period of 25.5 days (p=0.02). Comparison of time to RT-PCR negativity for males aged 6 to 15 years with other groups were not significant (all p>0.05).

**Figure 3:**
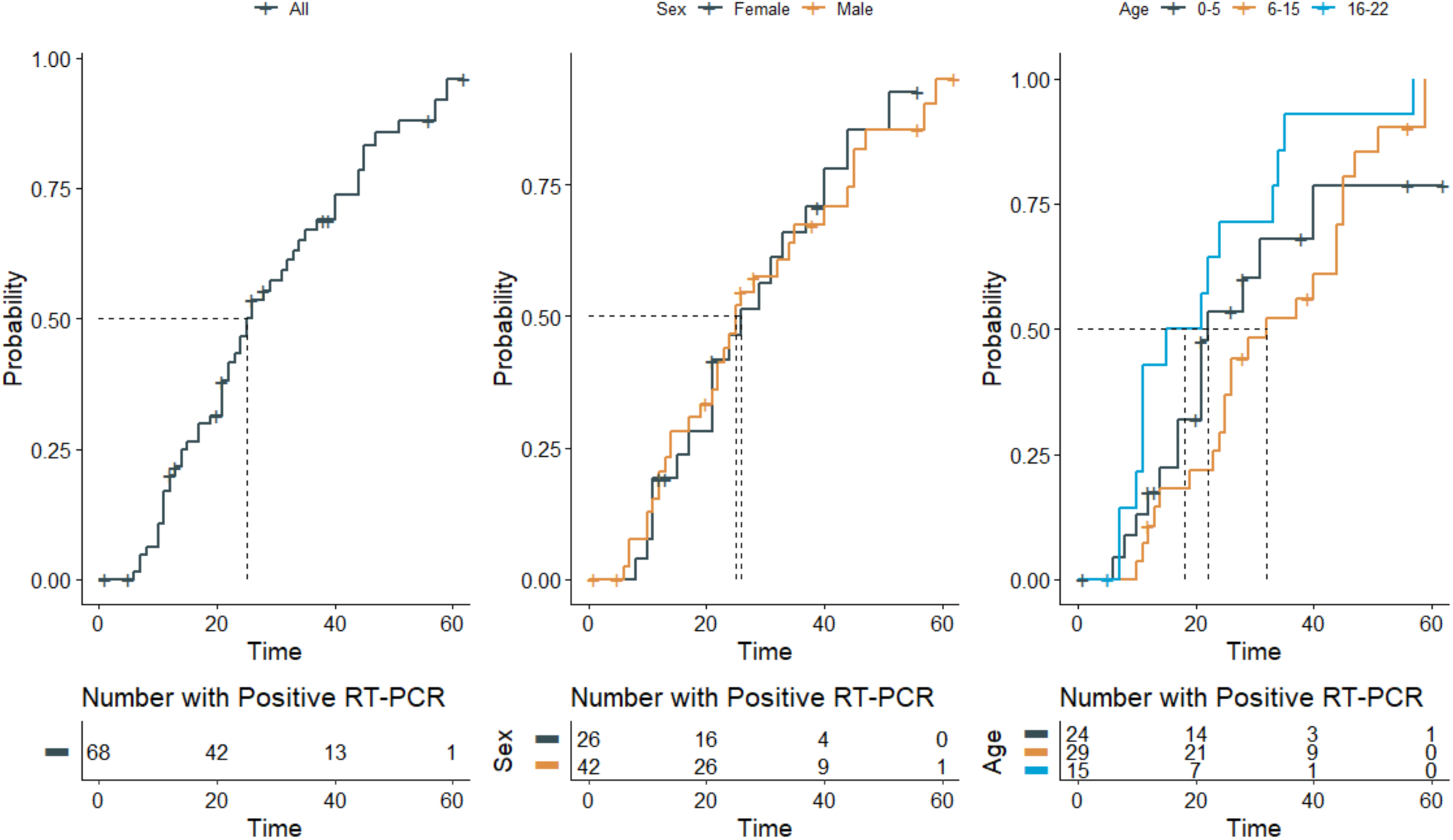
Time-to-event curves for RT-PCR positivity to negativity

**Figure 4:**
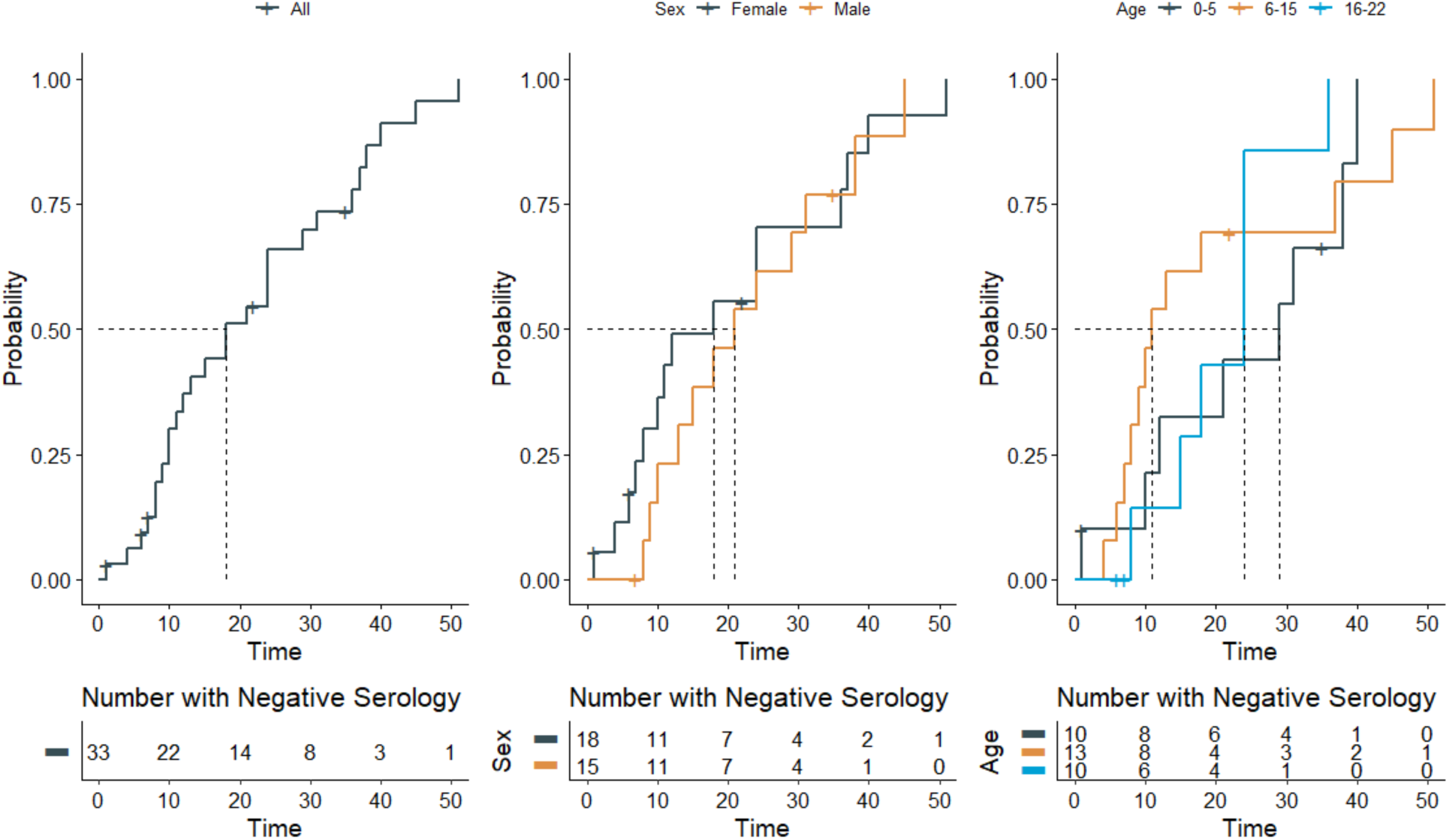
Time-to-event curves for RT-PCR positivity to seropositivity (Anti-SARS-CoV-2 IgG antibody ≥ 15 AU/mL)

The median time to seropositivity from RT-PCR positivity was 18 days (95% CI=12-31) (Figure 4a). No difference in time to seropositivity was found between females (median = 18 days) and males (median = 21 days) (χ^2^=0.8, p=0.4) (Figure 4b). The median number of days for seroconversion from initial RT-PCR positivity was 29 days for the 0 to 5 year group, 11 days for the 6 to 15 year group, and 24 days for the 16 to 22 year group. However, overall comparison of age groups did not demonstrate a significant difference (χ^2^=1.6, p=0.4) (Figure 4c). After adjustment for sex, the various age groups also did not demonstrate significant differences in time to seropositivity (χ^2^=0.6, p=0.7). Only 17 of 33 patients demonstrated antibody levels ≥ 80 AU/mL. The median time to reach adequate levels of neutralizing antibodies as defined by the manufacturer and this study was 36 days (95% CI=18-NA) (Figure 5a). No significance was found for sex (χ^2^=1.1, p=0.3) (Figure 5b), age (χ^2^=0.9, p=0.6), or age stratified for sex (χ^2^=1.7, p=0.4) (Figure 5c).

**Figure 5:**
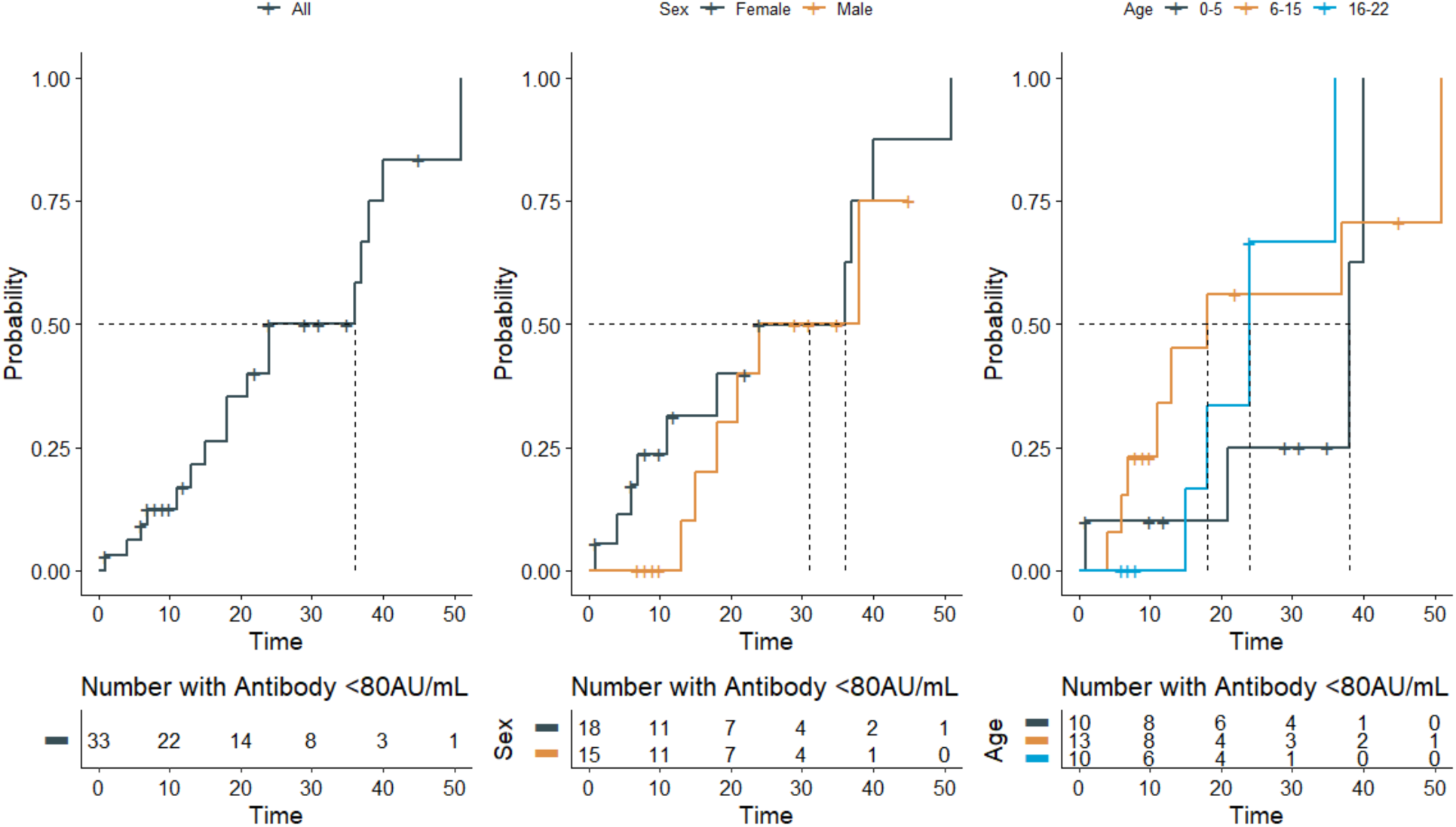
Time-to-event curves for RT-PCR positivity to reach neutralizing antibody levels (Anti-SARS-CoV-2 IgG antibody ≥ 80 AU/mL)

## Discussion

In this study, we demonstrated that IgG class antibodies directed against S1 and S2 glycoproteins could be detected in blood samples of children before viral clearance. Previous studies revealed that antibodies bound to the RBD epitope of SARS-CoV-2’s S1 glycoprotein are able to disrupt the virus-ACE2 interaction, thus blocking viral entry into human cells and enabling neutralizing capacity to the antibodies.^6^ While the RBD is located on the S1 subunit, the S2 subunit plays a crucial role in membrane fusion of the virus by conformation changes.^18, 19^ It was previously hypothesized for SARS-CoV that multiple antibodies targeting different epitopes might act synergistically.^20, 21^ As noted earlier, the antibody detection assay utilized in this study does not measure only antibodies targeting RBD but includes all antibodies to epitopes on S1 and S2 glycoproteins. We believe that this assay design may be beneficial in assessing antibody response in individuals with a polyclonal immune response to both S1 and S2 antigens with synergistic anti-viral activity.

Most of the SARS-CoV-2 literature related to viral kinetics focuses on moderate to severe adult cases of COVID-19. Han *et al*. observed 12 children with mild or asymptomatic COVID-19 and reported gradual viral load decrease in nasopharyngeal samples from 100% to 55% positivity over three weeks, ^22^ a finding which is comparable to our median period of 25 days for achieving nasopharyngeal RT-PCR negativity. Fafi-Kremer *et al*. reported immunological responses of 160 health care workers with mild or subclinical COVID-19 and demonstrated that even mild cases were usually, but not always, characterized by formation of neutralizing antibodies with an increase in neutralization activity over time.^23^ The authors reported that the variables associated with high neutralizing activity in a multivariate model included time to blood sampling from symptom onset, high BMI and male sex.^23^ We demonstrated that females between the ages of 6 to 15 years required a longer period for viral clearance when compared to other groups. This may be due to the age-dependent expression of ACE2 in the nasal epithelium, as demonstrated by Bunyavanich *et al*., where ACE2 gene expression was found to be significantly higher in older children (10 to 17 years) compared to younger children (< 10 years).^24^ Furthermore, it was suggested that gonadal hormones play a role in ACE2 expression and function.^25^ Taken together, increased duration of SARS-CoV-2 in the nasopharyngeal area could be caused by hormonal changes in adolescent females in this age group. As noted by Chun *et al*,^26^ different sections of the airway feature variable expression of ACE2, and prolonged presence of the viral genome in the upper respiratory tract may not correlate with the severity of COVID-19.

A strength of our study was the inclusion of patients from multiple pediatric age groups with sequential PCR testing, which allowed comparison between age groups and sex. One patient still demonstrated RT-PCR positivity 62 days after the initial positive result. Results of serial testing in pediatric patients have only been rarely reported in the literature to date. In a case series of 50 children, only four had repeat testing performed, and one patient demonstrated continued positivity 27 days after initial RT-PCR testing.^27^ In the adult population, retrospective evaluation of 191 adult patients from Wuhan reported the longest duration of viral shedding to be 37 days with a median of 20 days.^28^ It should be noted that detection of viral particles by molecular testing may not correlate with viable virus and transmissibility.^29^

For COVID-19, antibody competition with ACE2, the intended target of SARS-CoV-2, and binding affinity of the antibody for RBD are critical to neutralization of the virus.^6^ We demonstrated that the virus can still be detected in nasopharyngeal samples with low levels of circulating antibody but it becomes undetectable when levels reach neutralizing levels. This suggests that quantitative antibody results may be more useful for clinical management of patient. The U.S. Food and Drug Administration (FDA) has granted Emergency Use Authorization (EUA) to multiple viral and serologic tests for diagnosis and management of COVID-19.^15^ However, serologic assays for SARS-CoV-2 are still in early phases of development. As of July 26, 2020, no commercial assay was approved by FDA for quantitative reporting of the results.^15^ In the present study, we showed that time to reach a quantitative result corresponding to an antibody titer of 160 was associated with time to viral clearance. This titer level has also been recommended by the FDA to identify potential convalescent plasma donors. However, it has been observed that in the setting of other viral infections that seropositivity or antibody response may not correlate to immunity to the virus and that disease progression or reinfection is still possible. Tang *et al*. reported that humoral immunity mounted against SARS-CoV gradually decreased over time and disappeared due to the lack of peripheral memory B cell response in most individuals.^30^

## Limitations

This study has a number of limitations which include its retrospective nature and timing of viral and antibody testing being at the discretion of the ordering provider rather than at defined time intervals. We have not included symptom onset in our analysis since this project was solely based on laboratory data evaluation. In addition, the serologic assay used in the study had 94.3% positive and 100% negative agreement with a comparative enzyme-linked immunosorbent assay. Given the low optimal positive agreement, some false negative test results are expected.

## Conclusions

In summary, given the significant volume of testing performed, we provide a timeline of viral clearance and humoral response to COVID-19 in pediatric patients with additional comparisons amongst age groups and sex. We demonstrated that females between 6 to 15 years of age experienced longer persistence of viral genome in nasopharyngeal samples. It should be noted that presence of viral genome may not correlate with transmissibility. Antibodies were detectable in low titer preceding viral clearance. The timing of antibodies reaching titers that correlate with potentially neutralizing levels coincided with RT-PCR negativity in nasopharyngeal samples within a 24 to 25 day period after initial RT-PCR positivity. However, only approximately 50% (17 of 33 patients) actually achieved this antibody level at some point in their disease course.

## Data Availability

De-identified data would be provided upon request after approval from institutional approval.

## Acknowledgements

Authors would like to thank Eric Freeman and Celia Grant, MT(ASCP) for their efforts in this study.

---

**Supplemental Figure S1:**
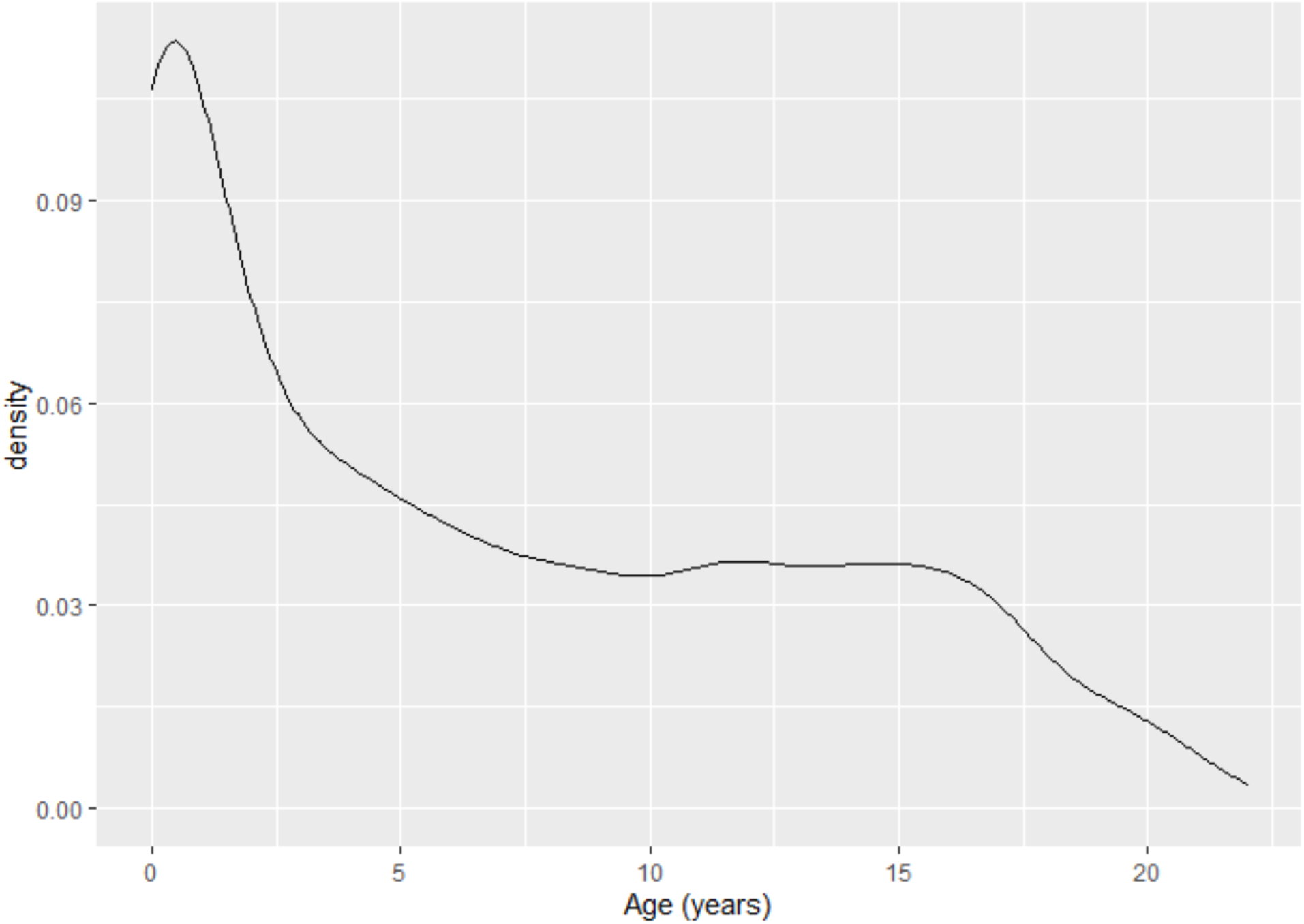
Age distribution of the patients underwent molecular and serology testing

